# Broad immunogenicity to prior SARS-CoV-2 strains and JN.1 variant elicited by XBB.1.5 vaccination in nursing home residents

**DOI:** 10.1101/2024.03.21.24303684

**Authors:** Yasin Abul, Clare Nugent, Igor Vishnepolskiy, Tiffany Wallace, Evan Dickerson, Laurel Holland, Iva Esparza, Mandi Winkis, Kazi Tanvee Wali, Philip A. Chan, Rosa R. Baier, Amy Recker, Matthew Kaczynski, Shreya Kamojjala, Alexander Pralea, Hailee Rice, Olubunmi Osias, Oladayo A. Oyebanji, Olajide Olagunju, Yi Cao, Chia Jung Li, Alex Roederer, Walther M. Pfeifer, Christopher L. King, Jurgen Bosch, Aman Nanda, Lynn McNicoll, Nadia Mujahid, Sakeena Raza, Rohit Tyagi, Brigid M. Wilson, Elizabeth M. White, David H. Canaday, Stefan Gravenstein, Alejandro B. Balazs

## Abstract

**Background:** SARS-CoV-2 vaccination has reduced hospitalization and mortality for nursing home residents (NHRs). However, emerging variants coupled with waning immunity, immunosenescence, and variability of vaccine efficacy undermine vaccine effectiveness. We therefore need to update our understanding of the immunogenicity of the most recent XBB.1.5 monovalent vaccine to variant strains among NHRs.

**Methods:** The current study focuses on a subset of participants from a longitudinal study of consented NHRs and HCWs who have received serial blood draws to assess immunogenicity with each SARS-CoV-2 mRNA vaccine dose. We report data on participants who received the XBB.1.5 monovalent vaccine after FDA approval in Fall 2023. NHRs were classified based on whether they had an interval SARS-CoV-2 infection between their first bivalent vaccine dose and their XBB.1.5 monovalent vaccination.

**Results:** The sample included 61 NHRs [median age 76 (IQR 68-86), 51% female] and 28 HCWs [median age 45 (IQR 31-58), 46% female). Following XBB.1.5 monovalent vaccination, there was a robust geometric mean fold rise (GMFR) in XBB.1.5-specific neutralizing antibody titers of 17.3 (95% confidence interval [CI] 9.3, 32.4) and 11.3 (95% CI 5, 25.4) in NHRs with and without interval infection, respectively. The GMFR in HCWs was 13.6 (95% CI 8.4,22). Similarly, we noted a robust GMFR in JN.1-specific neutralizing antibody titers of 14.9 (95% CI 7.9, 28) and 6.5 (95% CI 3.3, 13.1) among NHRs with and without interval infection, and a GMFR of 11.4 (95% CI 6.2, 20.9) in HCWs. NHRs with interval SARS-CoV-2 infection had higher neutralizing antibody titers across all analyzed strains following XBB.1.5 monovalent vaccination, compared to NHRs without interval infection.

**Conclusion:** The XBB.1.5 monovalent vaccine significantly elevates Omicron-specific neutralizing antibody titers to XBB.1.5 and JN.1 strains in both NHRs and HCWs. This response was more pronounced in individuals known to be infected with SARS-CoV-2 since bivalent vaccination.

**Impact Statement:** All authors certify that this work entitled “*Broad immunogenicity to prior strains and JN.1 variant elicited by XBB.1.5 vaccination in nursing home residents*” is novel. It shows that the XBB.1.5 monovalent vaccine significantly elevates Omicron-specific neutralizing antibody titers in both nursing home residents and healthcare workers to XBB and BA.28.6/JN.1 strains. This work is important since JN.1 increased from less than 0.1% to 94% of COVID-19 cases from October 2023 to February 2024 in the US. This information is timely given the CDC’s latest recommendation that adults age 65 and older receive a Spring 2024 XBB booster. Since the XBB.1.5 monovalent vaccine produces compelling immunogenicity to the most prevalent circulating JN.1 strain in nursing home residents, our findings add important support and rationale to encourage vaccine uptake.

**Key Points:**

- Emerging SARS-CoV-2 variants together with waning immunity, immunosenescence, and variable vaccine efficacy reduce SARS-CoV-2 vaccine effectiveness in nursing home residents.
- XBB.1.5 monovalent vaccination elicited robust response in both XBB.1.5 and JN.1 neutralizing antibodies in nursing home residents and healthcare workers, although the absolute titers to JN.1 were less than titers to XBB.1.5
- Why does this paper matter? Among nursing home residents, the XBB.1.5 monovalent SARS-CoV-2 vaccine produces compelling immunogenicity to the JN.1 strain, which represents 94% of all COVID-19 cases in the U.S. as of February 2024.

## Introduction

Severe acute respiratory syndrome coronavirus 2 (SARS-CoV-2) remains the most consequential respiratory virus in broad circulation.^1^ As of March 2024, SARS-CoV-2 has infected over 1.9 million NHRs and 1.8 million nursing home staff in the U.S., contributing to at least 171,000 resident and 3,000 staff deaths^2^. Nursing home residents (NHRs) are at particularly high risk for severe disease and adverse health outcomes. The burden and risks of disease are also important for the healthcare workers (HCW) who provide care in these settings. Unlike early in the pandemic, when SARS-CoV-2 vaccine coverage exceeded 50% for both staff and residents, only 7% of NH staff and 42% of NHRs have received the most recent XBB 1.5 monovalent vaccine.^2^ The evolution of highly immune evasive variants of SARS-CoV-2, waning immunity, immunosenescence, and variations in vaccine efficacy may compromise vaccine effectiveness in NHRs.

The lineage BA.2.86, first identified in August 2023, evolved into its currently circulating descendant, JN.1(BA.2.86.1.1) first identified in September 2023.^3,4^ JN.1 has more than 30 mutations compared to XBB.^5^ Both XBB and JN.1 variants, descendants from the Omicron BA.2 line, have amino acid substitutions that might augment escape from neutralizing antibodies. In the US, from the end of October, when JN.1 made up less than 0.1% of COVID-19 cases to March, 2024, JN.1 had spread to account for 98% of all COVID-19 cases.^6^ Despite common ancestry with the BA.2, the XBB lineage differs substantially from JN.1, that would raise reasonable concern about the ability of an XBB-based vaccine to provide cross-protective immunity to JN.1.^6^ Our understanding of the durability and nature of antibody protection against SARS-CoV-2 infection is still evolving. In the nursing home population, prior work has demonstrated a waning of SARS-CoV-2 mRNA vaccine immunity 3 to 6 months following each vaccine dose.^7–9^ The first and second monovalent boost and bivalent SARS-CoV-2 mRNA vaccine augmented immunity and demonstrated significant reduction in risk of infection, hospitalization and death at four months following vaccine administration.^10,11^ The newer monovalent XBB.1.5 vaccine has demonstrated adequate immunogenicity and cross-reactivity with current circulating strains in healthier, younger populations,^12–14^ but it is unknown whether this broader protection is also afforded to NHRs. We present an assessment of the immune response following XBB.1.5 monovalent vaccination to prior SARS-CoV-2 strains and the JN.1 variant among NHRs and a reference population of HCWs.

## Methods

### Ethical approval

This study was approved by the WCG Institutional Review Board. All participants or their legally-authorized representatives provided informed consent.

### Study design and participants

The current study is part of a longitudinal cohort study that has monitored immunological response to each SARS-CoV-2 mRNA vaccine dose in consented NHRs in Ohio and Rhode Island since SARS-CoV-2 vaccination began in December 2020.^7–9,11,15^ We have measured neutralizing antibody titers on serum samples collected from 447 unique NHR over the study period. Participants are sampled roughly 2-4 weeks before and after each vaccine dose and at 3-6 month intervals. ^7–9^ Not all study participants are sampled at each timepoint.

In this report, we focus on a subset of 61 NHR study participants who received both bivalent and XBB.1.5. monovalent vaccine doses. We additionally collected serum samples on 28 HCWs in Ohio as a reference group. XBB.1.5 vaccine administration started in September 2023 following FDA approval. Post-XBB.1.5. samples were collected 2-4 weeks after XBB.1.5 monovalent vaccine administration.

We analyzed NHR serum samples tested for neutralizing antibody titers against multiple SARS-CoV-2 strains, including ancestral Wuhan, Omicron BA.4/5, XBB.1.5, JN.1 using bead-based ELISA methods and pseudovirus neutralization assays, as detailed elsewhere.^7,8^

For data points related to bivalent vaccination and earlier vaccine doses, we defined a NHR as having had a “prior SARS-CoV-2 infection” at the time of each collected sample based on clinical criteria (any prior positive polymerase chain reaction or antigen test documented in the medical chart) or lab criteria (a significant increase in SARS-CoV-2 antibody levels, i.e. a rise outside of lab variance of anti-spike (S), N, or neutralizing assay results, that could not be explained by vaccination). We defined participants as “infection naive” at each time point if they did not meet the above criteria for prior infection.^7–9^ For data points related to the XBB.1.5 monovalent vaccine, we further classified NHR as having had an “interval SARS-CoV-2 infection” if they had a new SARS-CoV-2 infection between their prior bivalent vaccine dose and their XBB.1.5 monovalent vaccine dose. Interval infection was determined based on the same clinical and lab criteria described above. We did not stratify HCWs in this way because infection history was not available for the majority of HCWs with post-XBB.1.5 draws.

### Statistical analysis

We analyzed NHRs and HCW results separately. Within the longitudinal NHR cohort, we summarized the distributions of neutralizing titers over all vaccine doses for Wuhan and Omicron BA.4/5 strains, stratifying results at each sample time by infection naive vs. prior. Within the cohort of NHRs and HCW sampled following XBB.1.5 vaccine, we assessed the geometric mean titers with 95% confidence interval before and after XBB.1.5 monovalent vaccination for all strains and calculated the geometric mean fold rise (GMFR) from pre- to post-XBB.1.5 monovalent vaccine. We performed t-tests on log transformed titer fold change comparing the observed fold rise to a null value of 1. For the fold change analysis, we required participants to have pre-XBB.1.5. samples collected at least six months after their prior vaccine dose and excluded participants who received a second bivalent vaccine.

We provide all p values without adjustment. All analyses were performed in R (version 4.2.2).

## Results

Sample characteristics are presented in **Table 1**. The longitudinal cohort included 447 NHRs [median age 75 (IQR 69-86, 49% female,19% Black). The subset of study participants sampled following the XBB.1.5 monovalent booster included 61 NHRs [median age 76 (IQR 68-86), 51% female, 20% Black], and 28 HCWs [median age 45 (IQR 31-58), 46% female)] (**Supplementary Figure 1**).

**Table 1:** Sample characteristics

|  | Longitudinal<br>cohort of NHRs | NHRs sampled after<br>XBB.1.5 monovalent<br>vaccine | HCWs sampled after<br>XBB.1.5 monovalent<br>vaccine |
| --- | --- | --- | --- |
| Subjects, n | 447 | 61 | 28 |
| Age at consent,<br>median, (IQR) | 75<br>(69,86) | 75<br>(66,84) | 55<br>(25,58) |
| Age at consent,<br>Range | 33-104 | 48-104 | 21-64 |
| Male, n (%) | 229 (51%) | 30 (49%) | 15 (54%) |
| Female, n (%) | 218 (49%) | 31 (51%) | 13 (46%) |
| White, n (%) | 357 (80%) | 47 (77%) | 22 (79%) |
| Black, n (%) | 83 (19%) | 12 (20%) | 1 (4%) |
| Hispanic, n (%) | 3 (1%) | 0 | 1 (4%) |
| Asian, n (%) | 2(<1%) | 0 | 4 (14%) |
| Other race or<br>ethnicity, n (%) | 2(<1%) | 2 (3%) | 0 |
| Interval<br>Infection n (%) | 98 (22%) | 42 (69%) | NA |

*Notes.* Race and ethnicity unavailable for healthcare workers. HCW, health care worker; NHR, nursing home resident

**Figure 1** presents neutralizing antibody levels against Wuhan and BA.4/5 for the overall longitudinal cohort of NHRs from before the primary SARS-CoV-2 vaccine series until after XBB.1.5 vaccination; BA.4/5 titers were first measured beginning 9 months after the primary series was completed. Over the three months following vaccination with both monovalent and bivalent vaccinations, neutralizing antibody levels fell to Wuhan and BA.4/5, and, absent intercurrent infection or additional boosting, continued to wane through to 12 months post-vaccination. Both the bivalent and XBB.1.5 monovalent vaccinations, at least initially, boosted neutralizing antibody titers against Wuhan and BA.4/5. At virtually all time points, NHRs with prior SARS-CoV-2 infections had higher neutralizing antibody titers after each vaccine administration dose than infection-naive NHRs.

Within the NHR cohort sampled following XBB.1.5 monovalent vaccination, we noticed waning neutralizing antibody levels for Wuhan and BA.4/5 strains over the three-, six- and 12-month time points after bivalent vaccination in NHRs (**Figure 2**). NHRs with and without interval SARS-CoV-2 infection had robust GMFR [17.3 (95% CI 9.3, 32.4) and 11.3 (95% CI 5, 25.4) respectively] in XBB.1.5 subvariant specific neutralizing antibody titer levels following the XBB.1.5 monovalent vaccination (p<0.001). The XBB.1.5 monovalent dose also induced substantial rises in neutralizing antibodies not only in NHRs but also in HCWs by GMFR of 13.6 (p<0.001) (**Table 2, Figure 3**). The GMFR after XBB.1.5 monovalent vaccination was notably higher in XBB.1.5-specific neutralizing antibody levels compared to those of Wuhan and BA.4/5 specific neutralizing antibody levels (11.3 and 17.3 in XBB.1.5 subvariant, 6.5 and 6.3 in BA.4/5 subvariant, and 3.7 and 3.4 in Wuhan ancestral strain, respectively) (**Table 2**). Comparing to XBB.1.5 titers among NHRs with and without interval SARS-CoV-2 infection, we noticed a similarly robust GMFR in JN.1 subvariant specific neutralizing antibody titer levels following the XBB.1.5 monovalent vaccination (p<0.001) [14.9 (95% CI 7.9, 28) and 6.5 (95% CI 3.3, 13.1) respectively, and 11.4 (95% CI 6.2, 20.9) in HCWs] (**Table 2, Figure 3).**

**Figure 1.**
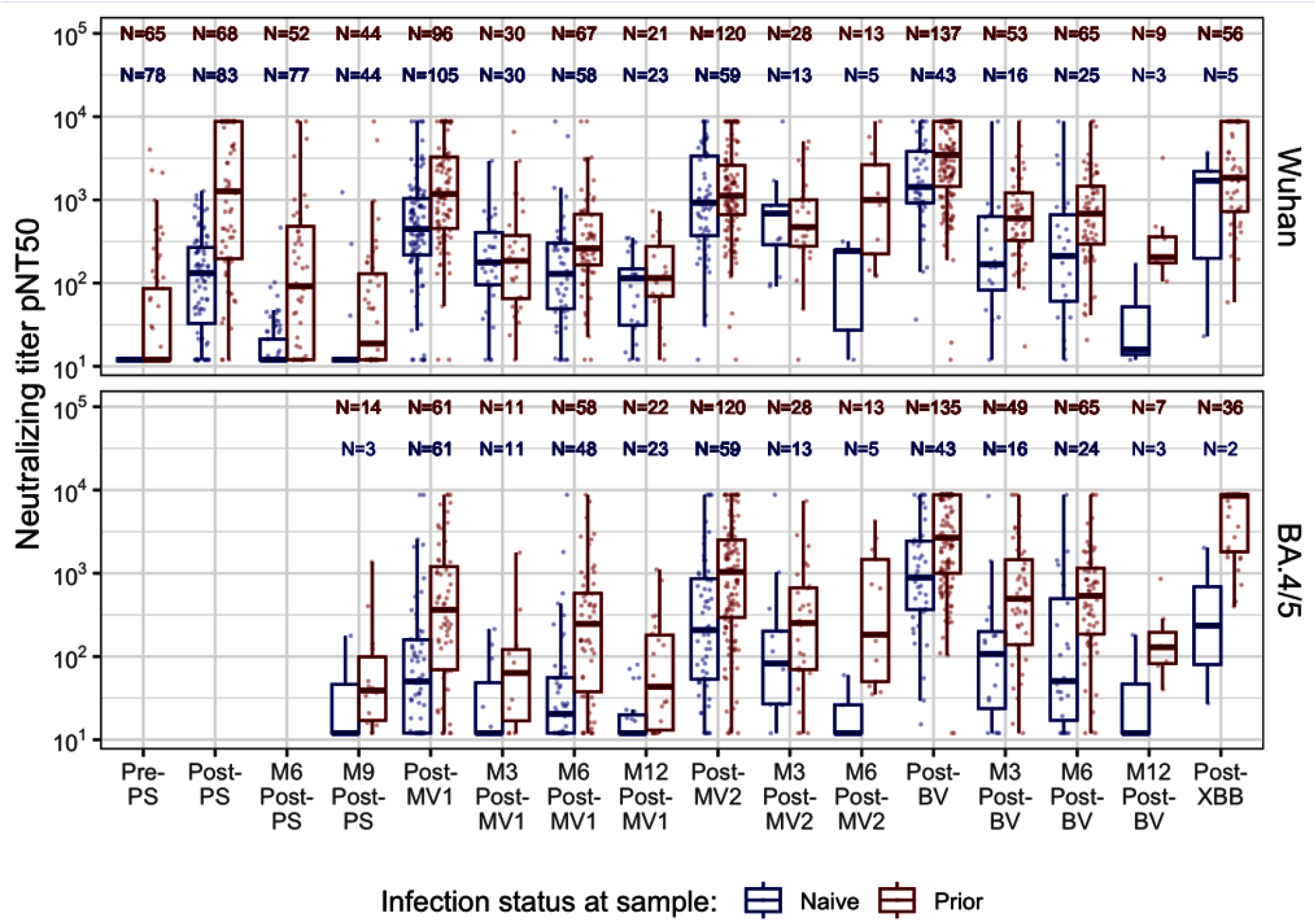
Neutralizing antibody titers against Wuhan and BA.4/5 in nursing home residents (n=447), from SARS-CoV-2 primary vaccine series through XBB.1.5 monovalent vaccination *Notes*. PS = Primary series; MV1 = first monovalent vaccine; MV2 = second monovalent vaccine; BV = bivalent vaccine; XBB = XBB monovalent vaccine; M3= month 3; M6 = month 6, M9 = month 9, M12 = month 12. In cases of SARS-CoV-2 infection, samples within subjects were excluded from the time of breakthrough infection until the next vaccine dose.

**Figure 2.**
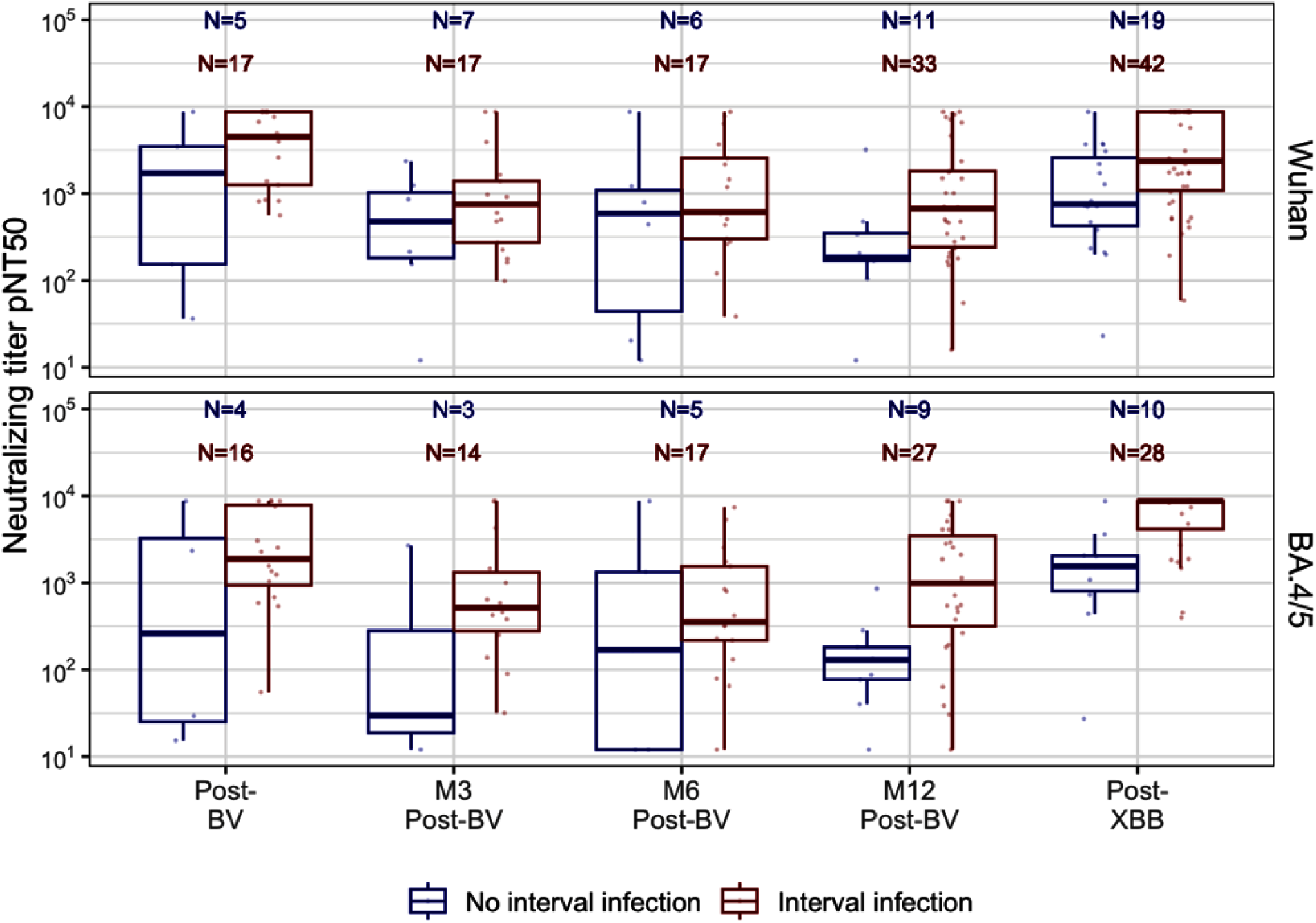
Neutralization titers against Wuhan and BA.4/5 after bivalent and XBB.1.5 monovalent SARS-CoV-2 mRNA vaccination, among nursing home residents with and without interval infection (n=61 *Notes*. Interval SARS-CoV-2 infection is defined as an infection occurring between a participant’s prior bivalent vaccine dose and the XBB.1.5 monovalent dose. BV = bivalent vaccine; XBB = XBB monovalent vaccine; M3 = month 3; M6 = month 6, M12 = month 12. Post- BV and Post-XBB samples were collected 2-4 weeks after vaccine dose. *Healthcare workers’ all prior infection status combined as insufficient data available to categorize them

**Figure 3.**
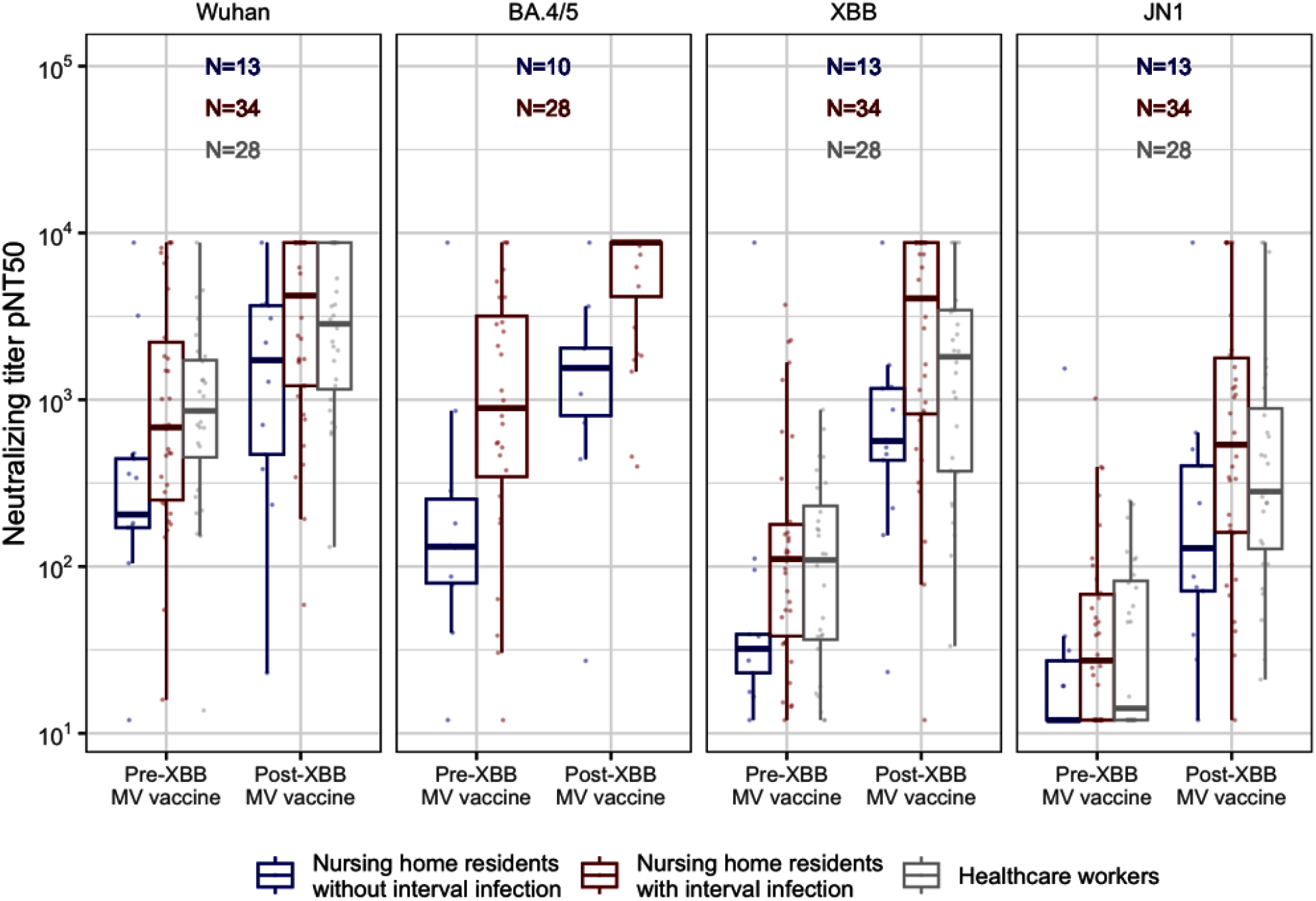
Neutralization titers against Wuhan, BA.4/5, XBB and JN1 after XBB.1.5 monovalent SARS-CoV-2 mRNA vaccination among NHRs and HCWs with and without interval infection *Notes*. Interval SARS-CoV-2 infection is defined as an infection occurring between a participant’s prior bivalent vaccine dose and the XBB.1.5 monovalent dose. MV = monovalent vaccine; post-XBB samples were collected 2-4 weeks after vaccine dose.

**Table 2:** Wuhan, BA.4/5 and XBB.1.5 neutralizing antibody response in nursing home residents and health workers with and without interval SARS-CoV-2 infection since bivalent vaccination

| Assay<br>Neut | Group | n | Pre-XBB.1.5<br>GMT(CI) | Post-XBB.1.5<br>GMT(CI) | GMFR<br>(CI) | p-value |
| --- | --- | --- | --- | --- | --- | --- |
| Wu | HCW* | 28 | 783<br>(473, 1295) | 2513<br>(1640, 3851) | 3.2<br>(2.1, 4.9) | <0.001 |
| Wu | NHRs<sup>uninfected</sup> | 13 | 302<br>(116, 786) | 1124<br>(433, 2916) | 3.7<br>(1.9, 7.1) | <0.001 |
| Wu | NHRs<sup>infected</sup> | 34 | 807<br>(463, 1409) | 2719<br>(1698, 4355) | 3.4<br>(1.8, 6.2) | <0.001 |
| BA.4/5 | NHRs<sup>uninfected</sup> | 10 | 171<br>(48, 614) | 1115<br>(370, 3361) | 6.5<br>(2.5, 16.8) | 0.002 |
| BA.4/5 | NHRs<sup>infected</sup> | 28 | 823<br>(410, 1652) | 5145<br>(3601, 7350) | 6.3<br>(3.3, 11.7) | <0.001 |
| XBB | HCW* | 28 | 88<br>(53, 147) | 1198<br>(673, 2132) | 13.6<br>(8.4, 22) | <0.001 |
| XBB | NHRs<sup>uninfected</sup> | 13 | 49<br>(18, 136) | 555<br>(242, 1275) | 11.3<br>(5, 25.4) | <0.001 |
| XBB | NHRs<sup>infected</sup> | 34 | 121<br>(70, 208) | 2089<br>(1160, 3760) | 17.3<br>(9.3, 32.4) | <0.001 |
| JN.1 | HCW* | 28 | 32 (21, 49) | 365 (196, 677) | 11.4<br>(6.2, 20.9) | <0.001 |
| JN.1 | NHRs<sup>uninfected</sup> | 13 | 24<br>(11, 52) | 154<br>(56, 423) | 6.5<br>(3.3, 13.1) | <0.001 |
| JN.1 | NHRs<sup>infected</sup> | 34 | 37<br>(24, 57) | 552<br>(289, 1053) | 14.9<br>(7.9, 28) | <0.001 |

Notes. \*Interval infection status of HCWs unavailable.
NHRs<sup>uninfected</sup> : Nursing home residents without interval infection
NHRs<sup>infected</sup> : Nursing home residents with interval infection
CI: Confidence interval; GMT: Geometric mean titer; GMFR: Geometric mean fold rise; Wu: Wuhan

Within the subset XBB cohort of NHRs, we similarly observed higher neutralizing titers in those with an interval SARS-CoV-2 infection that persisted after XBB.1.5 monovalent vaccination (**Figure 3**).

## Discussion

This is the first study, to our knowledge, to demonstrate robust neutralizing antibody response for NHRs in the XBB.1.5 and JN.1 subvariant era. We report broad immunogenicity to variant strains by XBB.1.5 monovalent SARS-CoV-2 vaccines in both NHRs and HCWs. We find that both groups develop robust neutralizing antibody titers to XBB.1.5 monovalent vaccine, but that Wuhan and anti-omicron BA.4/5 neutralizing antibody titers decreased substantially over the three to 12 months following the first or second mRNA bivalent boosters. Those higher humoral immunity titers in the setting of hybrid immunity likely play an important role in increased protection in NHRs and HCWs. Our findings suggest the new XBB.1.5 monovalent vaccine likely augments the protection against SARS-CoV-2 infection caused by recently circulating XBB.1.5 subvariants in older vulnerable NHRs, may enhance immunity against variant strains, and may support ongoing efforts to prevent SARS-CoV-2 morbidity and mortality in NHRs.

Preliminary data from phase II-III trials indicate a boost in neutralizing antibody titers against both XBB.1.5 lineages and BA.2.86.^12^ In the most recent real-world data, Marking et al. (2024) reported a significant increase in neutralization activity against all variants tested after XBB.1.5 vaccination, with a more than 10-fold increase in GMT two weeks after vaccination (84 to 869) in 24 HCWs (median age = 64 years)^14^ and Stankov et al. (2024) reported a rise in GMT neutralization from 27 to 967 for XBB.1.5 and from 28 to 906 for XBB.1.16 after the updated monovalent Omicron XBB.1.5 vaccine (median age = 45).^13^ Our study cohort demonstrated 13.6 GMFR (from 88 to 1198) in neutralizing antibodies to XBB.1.5 in HCWs, 11.3 GMFR (from 49 to 555) in NH without known SARS-CoV-2 infection since bivalent vaccine dose, and 17.3 GMFR (from 121 to 2089) in NHRs with known interval infection. The current study provides the first data from NHRs (n=61, median age 76). Previously, Chalkias et al. (2023) reported similar GMFRs of neutralizing antibody levels in ancestral, BA.4/5 and XBB.1.5 sub variants after monovalent XBB.1.5 vaccine in a relatively younger study population (n=50, median age 55).^12^

We were encouraged by the finding that a similarly robust GMFR in JN.1 subvariant specific neutralizing antibody to that arising to the XBB.1.5 monovalent vaccination occurred both in NHRs and in HCWs, and both with and without interval infection. However, we also note that absolute JN.1 specific neutralizing antibody titers after XBB.1.5 monovalent vaccination were far below those of XBB specific neutralizing titers after XBB.1.5 monovalent vaccination. The most recent early estimates of XBB.1.5 monovalent vaccine effectiveness data while JN.1 was the prevailing circulating strain indicate 54% (95% CI:46-60%) protection against symptomatic SARS-CoV-2 infection compared to unvaccinated individuals.^11,16,17^

The protection against SARS-CoV-2 infection ^18^ and clinical illness has been reported as more durable than that produced by vaccination alone.^19^ However, previous studies found the most robust and persistent protection against SARS-CoV-2 infection is in individuals with hybrid immunity (i.e., immunity developed by the combination of both infection and vaccination).^20–24^ This study demonstrates that both hybrid immunity and receipt of the XBB.1.5 monovalent vaccine augmented the neutralizing antibody titers to a higher level, compared to individuals without SARS-CoV-2 infection, in the setting of the highly antibody-evasive variant of XBB.1.5. Future studies will need to determine the durability of hybrid immunity after XBB.1.5 monovalent vaccination and its determinants for potential longer durability in setting of newly evolving highly antibody evasive strains.

It has been suggested that hybrid immunity has helped protect against the highly antibody-evasive variants including XBB.1.5 and EG.5, and substantially contributes to the declining COVID-19 hospitalizations and mortality.^24^ We should take hybrid immunity into consideration for booster vaccination strategies since hybrid immunity is so common in the general population. However, NHRs continue to have an elevated risk of severe infection, given their high prevalence of immunosenescence and multiple morbidities, and the substantial exposure risk to infection conferred by the congregate nature of nursing home living environment. Therefore, NHRs have an opportunity for deriving added clinical benefit from boosting.

These study results are clinically important as neutralizing antibodies correlate with enhanced protection against symptomatic SARS-CoV-2 infection.^25,26^ Different studies conducted with various different SARS-CoV-2 strains reported variable cutoff levels of neutralizing antibodies to achieve protection against symptomatic SARS-CoV-2 infection.^26,27^ However, particularly in the current hybrid immunity era, T cell immunity might have a key role in both durability and protection of the vaccine besides the antibody response.^28^ McConeghy et. al. (2022) reported that vaccine effectiveness of 2nd monovalent mRNA booster dose against SARS-CoV-2 related hospital admission or mortality was 73.9% and 89.6% for death alone in comparison with single booster dose among NH individuals.^29^

Potential limitations of the current study include the small sample size, particularly for HCW participants which may limit generalizability in this group. We did not evaluate the vaccine induced T-cell immunity. While previous work has demonstrated that pseudovirus neutralization assays are comparable to assays performed with live virus, it is possible that this is no longer the case for more recent strains derived from B.2.86. Potential for selection bias is another limitation, since our study sample only includes participants who received the last vaccine and those who opted to receive it are not necessarily generalizable to NHRs and HCWs more broadly, who have low vaccine coverage in this setting 41% of NHRs and 7% of HCWs). The study also excludes those who got the second bivalent vaccine. A strength of this study is the longitudinal data from before the primary vaccine series through the XBB.1.5 monovalent booster, allowing for a comprehensive assessment of vaccine impact over time as new virus variants have emerged.

## Conclusion

This is the first study of XBB.1.5 and JN.1-specific neutralizing antibody responses after the XBB.1.5 monovalent vaccine in NHRs and shows comparable results among both NHRs and a comparison group of HCWs This is particularly important in the context of emerging variants coupled with waning immunity, immunosenescence, and variable vaccine effectiveness in NHRs and other older adults.

## Data Availability

All data produced in the present study are available upon reasonable request to the authors

## Conflict of Interest (Declaration of commercial interest)

S.G. and D.H.C. are recipients of investigator-initiated grants to their Universities from Pfizer, to study pneumococcal vaccines (S.G. and D.H.C.), Sanofi Pasteur and Seqirus, to study influenza vaccines (S.G. and D.H.C., and Genentech, to study influenza antivirals (S.G.). S. G. also consults for Seqirus, Sanofi, Merck, Vaxart, Novavax, Moderna and Janssen; has served on the speaker’s bureaus for Seqirus and Sanofi; and reports personal fees from Pfizer and data and safety monitoring board (DSMB) fees from Longevoron and SciClone.

## Author contributions

**Study concept and design:** SG, DHC, EMW, RB

**Acquisition of subjects and/or data**: CN, IV, IE, TW, ED, LH, HR, MK, KTW, AP, OOs, OAO, OO, AR, CJL, YC, AR, AN, LM, RT, SR, NM, ABB, SK, MW, WP

**Analysis and interpretation of data:** BW, OAO, CLK, ABB, DHC, SG, YA

**Preparation of manuscript:** YA, SG, DHC, EMW, BW, RB, ABB, PC, CLK

## Sponsor Roles

The sponsor had no role in the design, methods, data analysis or preparation of the manuscript.

## Supplemental materials

**Supplementary Figure 1:**
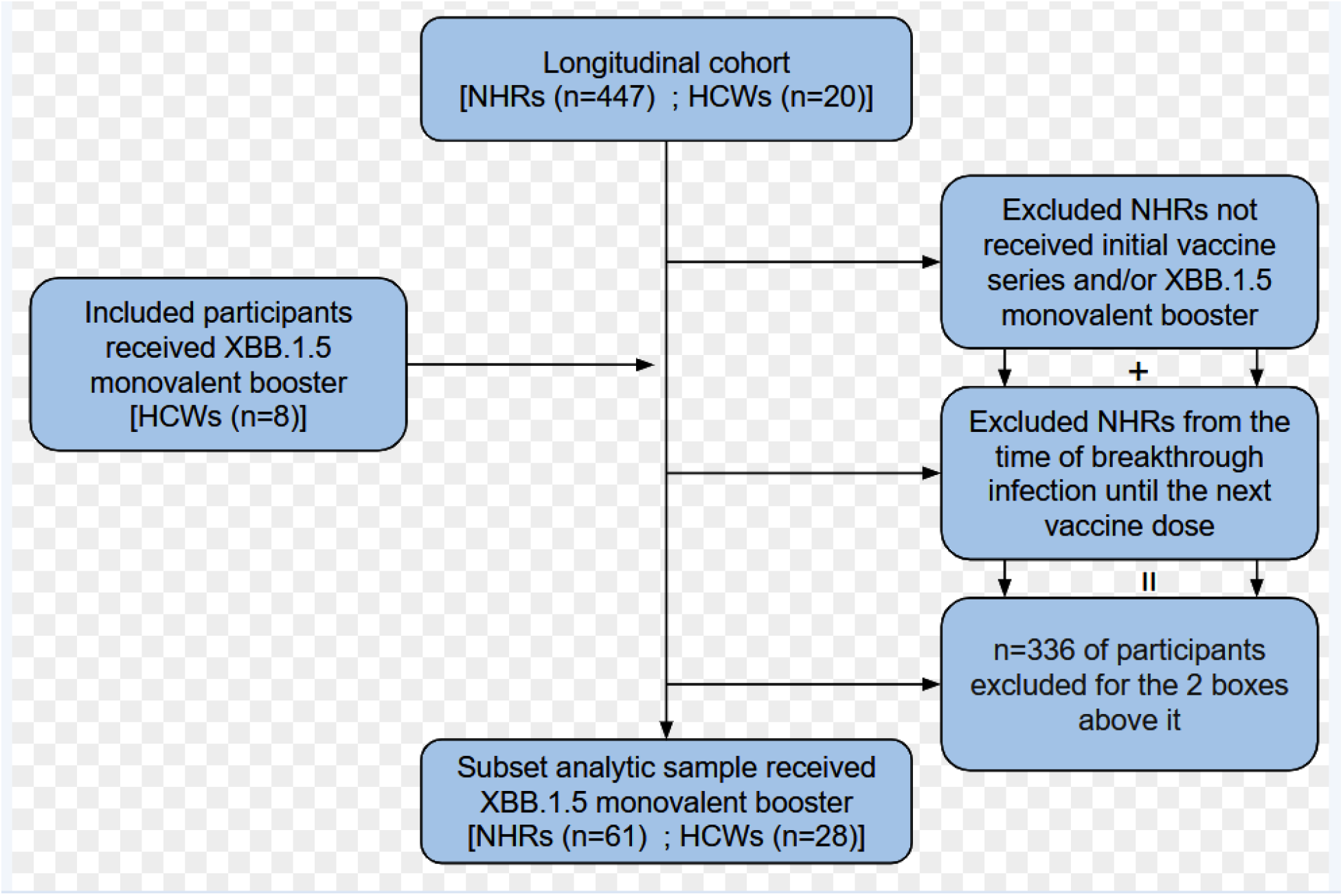
Consort diagram showing longitudinal cohort started to be recruited from before primary series of SARS-CoV-2 vaccination period till after XBB.1.5 monovalent vaccination period.

