## Supplementary figures and images for "Broad immunogenicity to prior SARS-CoV-2 strains and JN.1 variant elicited by XBB.1.5 vaccination in nursing home residents"

### Supplemental Figure 1

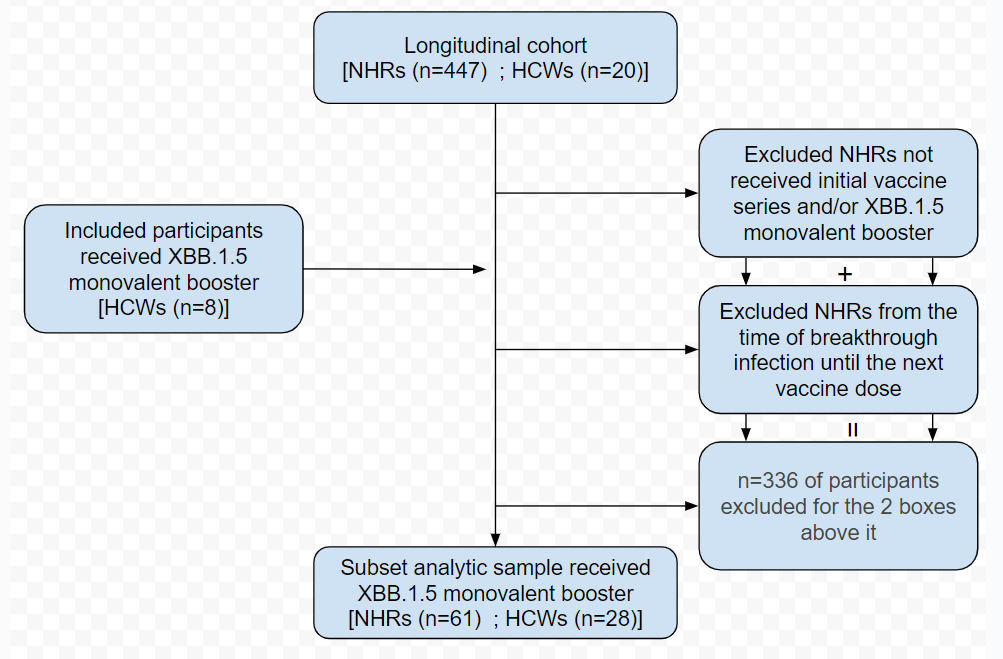
